# Multi-Omics Molecular Profiling Enables Rapid Diagnosis of Erythrodermic Skin Diseases

**DOI:** 10.1101/2025.09.12.25335624

**Authors:** Pia-Charlotte Stadler, Johannes B. Mueller-Reif, Katrin Kerl-French, Georg Wallmann, Lucas Diedrich, Laurie Eicher, Maximilian Zwiebel, Doris Helbig, Werner Kempf, Rudolf Stadler, Sonja Senner, Matthias Neulinger-Munoz, Mohammed Mitwalli, Anne-Sophie Boehm, Marius Winkler, Valerie Glatzel, Leonie H. Frommherz, Anna Leonhardt, Nora Aszodi-Pump, Benjamin Kendziora, Zeno Fiocco, Anna Oschmann, Michaela Maurer, Annika Sander, Julia Leding, Irma Kupf, Nina Janjic, Surina Frey, Sophia Czell, Benjamin M. Clanner-Engelshofen, Nicholas Moellhoff, Ruben A. Ferrer, Christiane Pfeiffer, Burkhard Summer, Eva M. Oppel, Felix Lauffer, Michael J. Flaig, Teodora Pumnea, Takashi K. Satoh, Matthias Mann, Lars E. French, Thierry M. Nordmann

**Affiliations:** Molecular and Spatial Biology of Skin, Max Planck Institute of Biochemistry, Martinsried, Germany; Department of Dermatology and Allergy, University Hospital, LMU Munich, Germany; Proteomics and Signal Transduction, Max Planck Institute of Biochemistry, Martinsried, Germany; Department of Dermatology, Thalkirchner Street Hospital, Munich Municipial Hospital Group, Munich, Germany; Department of Dermatology, University Hospital Zurich, Zurich, Switzerland; Faculty of Medicine, University of Zurich, Zurich, Switzerland; Department of Dermatology, University Hospital Cologne, Cologne, Germany; Kempf und Pfaltz Histologische Diagnostik, Zurich, Switzerland; University Clinic for Dermatology, Johannes Wesling Medical Center, UKRUB, University of Bochum, Minden, Germany; Division of Hand, Plastic and Aesthetic Surgery, University Hospital, LMU Munich, Germany; Dr. Phillip Frost Department of Dermatology & Cutaneous Surgery, University of Miami, Miller School of Medicine

## Abstract

Erythroderma is an acute and potentially life-threatening inflammatory condition characterized by redness and scaling of > 90% of the skin. Its treatment is challenging because various underlying skin diseases can cause erythroderma and are difficult to distinguish. Here, we performed in-depth proteomics and transcriptomics analyses of skin from 96 patients with erythroderma caused by five different diseases, including pityriasis rubra pilaris, psoriasis, atopic dermatitis, cutaneous T-cell lymphoma, and drug-induced maculopapular rash. High-throughput workflows enabled in-depth molecular profiling, identifying over 9,300 proteins and 17,200 protein coding transcripts, revealing distinct molecular signatures for each disease. The proteome showed elevated expression of type 2 immunity associated Charcot-Leyden crystal in skin of atopic dermatitis, potentially contributing to NLRP3-driven chronic inflammation in this disease. Complementary transcriptomic analysis demonstrated selective upregulation of IL17C in pityriasis rubra pilaris, strongly correlating with increased IL1 family cytokine expression. Interestingly, only a subset of these patients expressed this IL17C-IL1 signature, suggesting treatment-relevant disease endotypes. Through multi-omics integration, we uncovered disease-specific molecular signatures consistently altered at both protein and transcript levels. In particular, we identified elevated expression of T-cell regulator RASAL3 in cutaneous T-cell lymphoma, which has not been explored in its pathogenesis so far. To translate these molecular profiles into clinical utility, we expanded our adaptive machine-learning algorithm (ADAPT-Mx) for tissue based-disease classification. This achieved 76.6% diagnostic accuracy, substantially outperforming combined conventional clinical and histopathological methods (59.5%). This study provides a template for precision diagnostics in erythroderma and demonstrates the clinical potential of multi-omic profiling in severe inflammatory skin diseases.

## Introduction

Erythroderma, also known as exfoliative dermatitis, is a dermatological emergency characterized by erythema and scaling involving more than 90% of the body surface area (BSA) that is due to widespread inflammation of the skin and has an incidence of 0.001% (*1*). Erythroderma represents the most acute clinical form of different underlying inflammatory and malignant skin diseases. Most commonly, these include psoriasis (Pso), atopic dermatitis (AD), pityriasis rubra pilaris (PRP), cutaneous T-cell lymphoma (CTCL) and maculopapular drug reactions (MPR). Due to the uniform clinical presentation, identifying the underlying causal disease remains one of the greatest diagnostic challenges in modern dermatology (*2, 3*).

The current diagnostic approach relies on a combination of medical history, clinical examination, histopathology and clinical follow-up. However, there are inherent limitations to this approach, as histopathologic accuracy ranges from 48-66%, only a minority of patients have prior diagnosed causal skin-disease, and clinical follow-up which is sometimes required for final diagnosis delays the onset of specific therapy (*4*). As a result, misdiagnosis occurs frequently and time to diagnosis may take years (*5, 6*). This diagnostic uncertainty negatively impacts patient care, leading to inadequate treatment and further disease progression (*7, 8*). In addition, the acute and widespread inflammatory response causes severe systemic symptoms, and is associated with ∼ 9% mortality (*2*). Accordingly, there is an unmet need for improved diagnostic modalities.

Molecular omic profiling can substantially improve molecular classification of disease and targeted therapy thereof (*9, 10*). Increasingly compatible with routine clinical specimens, translational omic studies are proving particularly useful for rare diseases, where prospective profiling can take years (*11, 12*). Molecular profiling is particularly powerful to dissect the profoundly heterogeneous group of inflammatory skin diseases, aiming to achieve more accurate diagnosis and precision therapeutics (*13, 14*). To date, such studies have focused largely on single omic modalities, most commonly transcriptomics (*15–17*). In contrast, the potential value of systematic and unbiased analysis using mass spectrometry (MS)-based proteomics remains unexplored (*18*).

Here, we present the first comprehensive integration of high-resolution MS-based proteomics with transcriptomics in erythrodermic skin diseases. This approach enables molecular dissection of disease mechanisms at unprecedented depth and reveals diagnostic signatures inaccessible to conventional pathology. By combining these multi-omic profiles with an adaptive machine learning classifier, we establish a framework for accurate, individualized diagnosis of one of dermatology’s most challenging clinical presentations.

## Results

### High-throughput proteomics and transcriptomics of erythrodermic skin

We performed transcriptomic and proteomic profiling of erythrodermic (n = 96) and healthy skin (n = 20) from retrospectively collected paraffin-embedded, formalin-fixed (FFPE) tissue blocks. Two independent erythrodermic cohorts were assembled, including patients with pityriasis rubra pilaris (PRP; n = 19), psoriasis (Pso; n = 18), atopic dermatitis (AD; n = 20), cutaneous T-cell lymphoma (CTCL; n = 18) and maculopapular rash (MPR; n = 21; Fig. 1; Extended Data Fig. 2D). All cases were histologically validated prior to inclusion (Extended Data Fig. 1A). From each sample, sequential sections were then obtained for further proteomic and transcriptomic analysis. In parallel to the standardized transcriptomic workflow, we processed all samples for proteomic analysis in a semi-automated fashion, followed by measurement on the Evosep LC system coupled to the novel Astral mass spectrometer using narrow window data-independent acquisition (DIA) at 60 samples per day (*18–20*). This workflow enabled us to identify a cumulative ∼9,400 unique proteins across all patients and disease entities, spanning more than five orders of magnitude in abundance, including low-abundant interleukins (Extended Data Fig. 2B). Following data acquisition of the proteome and transcriptome from each sample, we performed comprehensive bioinformatic analysis to identify novel biological regulators across the disease entities (*21–23*). Ultimately, we leveraged our data in combination with machine learning algorithms to accurately predict the underlying disease (Fig. 1) (*24*).

**Fig. 1.**
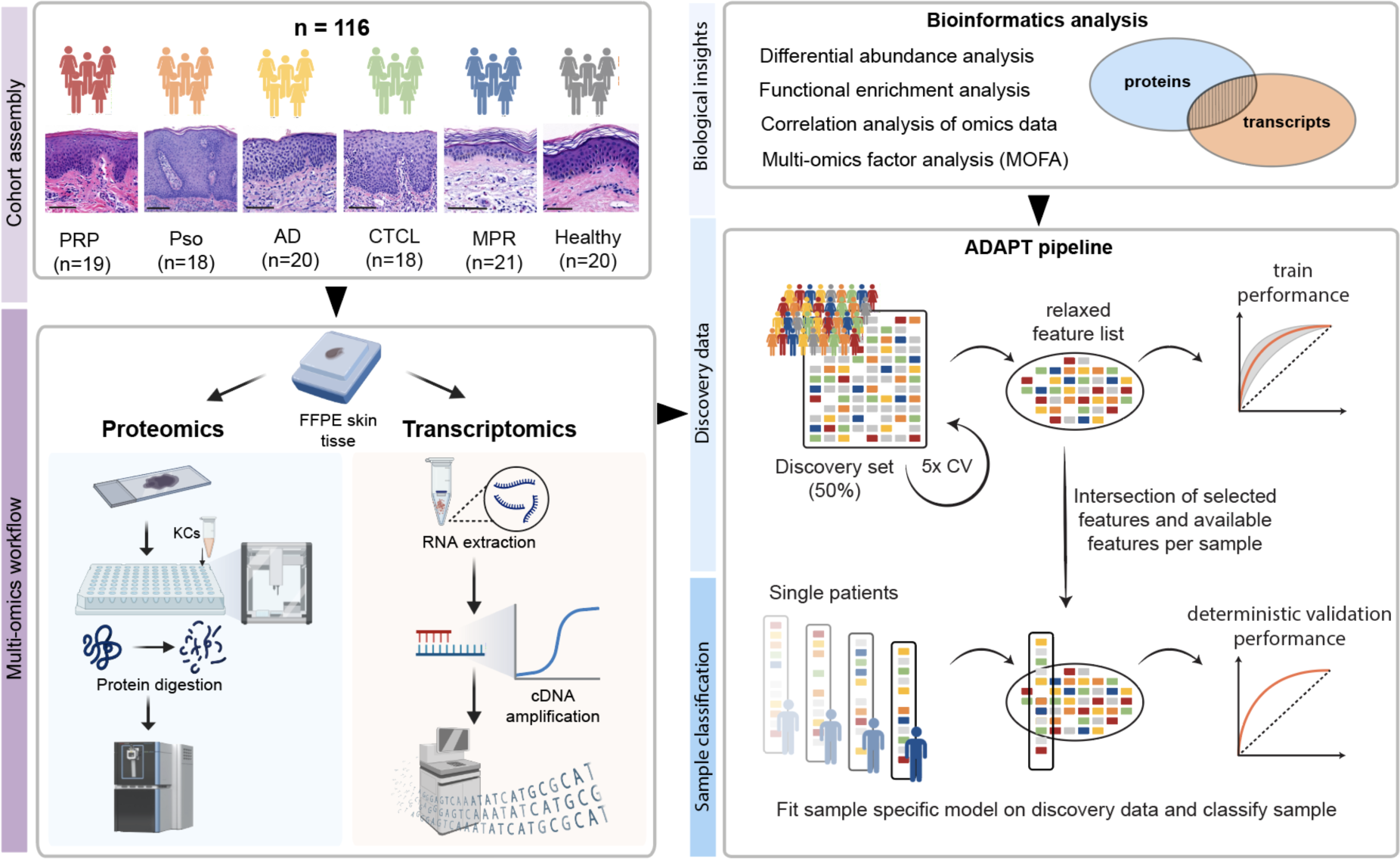
Workflow for multimodal analysis of erythrodermic skin. Cohort assembly (n = 116) of pityriasis rubra pilaris (PRP), psoriasis (Pso), atopic dermatitis (AD), cutaneous T-cell lymphoma (CTCL), maculopapular rash (MPR) and healthy controls. Sequential formalin-fixed and paraffin-embedded (FFPE) tissue sections were subjected to hematoxylin and eosin (H&E) staining, mass spectrometry-based proteomics and transcriptomics. Sample preparation was performed using a semi-automatic workflow for both modalities. After data acquisition, we performed biostatistical and multi-omics factor analysis (MOFA). Finally, we applied the ADAPT-Mx (Adaptive Diagnostic Architecture for Personalized Testing) pipeline that integrates molecular signatures from both omics modalities to enhance disease classification in erythroderma through adaptive, per-sample modeling. Partially created in BioRender.

### Disease-specific proteome of lesional skin in erythroderma

To identify disease-specific molecular mechanisms, we first analyzed the comprehensive proteome profiles generated across all patient cohorts. We detected ∼6,400 proteins on average per sample with high data completeness and reproducibility (Fig. 2A; Extended Data Fig. 2A-G). The proteome segregated the diseases, with psoriasis and PRP samples predominantly clustering together and CTCL and MPR forming a separate group (Extended Data Fig. 2H). This separation towards psoriasis/PRP was driven by proteins of the epidermal barrier (CDSN, SBSN, SERPINB7/12, RNASE7), while proteins involved in lymphocyte migration (DOCK2, HCLS1, MSN, SNX6) were important for CTCL/MPR (Extended Data Fig. 2I). Pairwise differential expression analysis identified numerous dysregulated proteins across all conditions, and the most pronounced changes were in comparison to healthy controls, including many ‘alarmins’ of keratinocyte inflammation (S100A7-9, KRT6A, KRT16; Fig. 2B; Extended Data Fig. 3B). Importantly, our proteomic data also revealed distinct molecular signatures between clinically related diseases (Fig. 2C-F; Extended Data Fig. 3A). PRP and psoriasis represent one such challenging diagnostic pair. In psoriasis, we observed higher expression of nicotinamide phosphoribosyltransferase (NAMPT), a key player in NF-κB signaling, as well as neutrophil-derived proteins transcobalamin (TCN1) and myeloperoxidase (MPO), consistent with the established role of neutrophils in psoriasis pathogenesis (Fig. 2C, D) (*25, 26*). In contrast, PRP showed elevated expression of skin barrier proteins (FLG, FLG2) and keratinocyte differentiation markers (KRT23; Fig. 2C), along with upregulated IL-1 signaling (Extended Data Fig. 2K), which aligns with reported clinical response to IL-1 antagonism in refractory PRP (*27*).

**Fig. 2.**
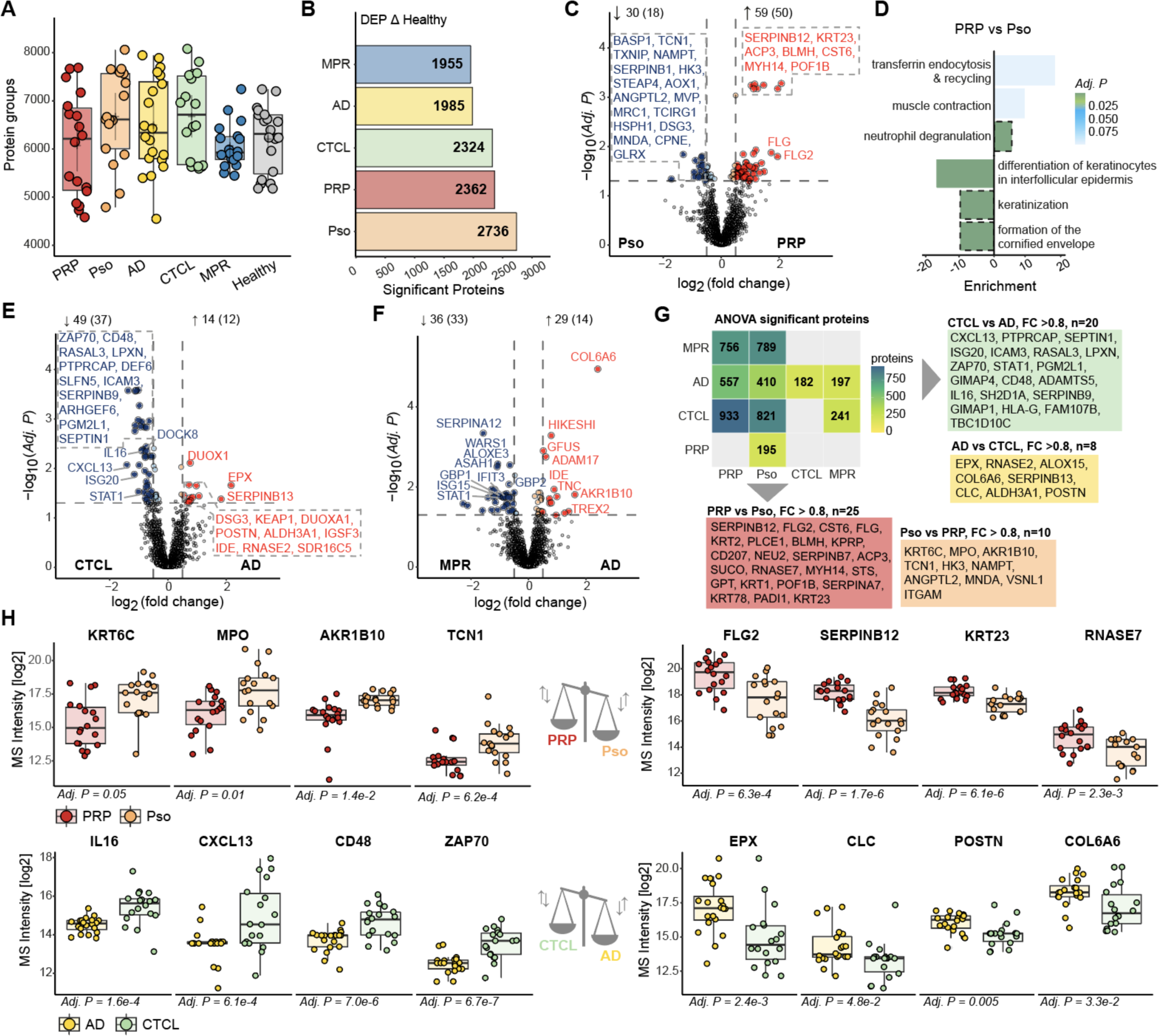
The proteome of erythrodermic skin diseases. **a.** Number of proteins identified across the cohorts (n= 113 individuals). **b.** Number of differentially expressed proteins (DEPs) for each disease versus healthy (n = 113 individuals). **c, e, f.** DEPs between psoriasis versus PRP (n = 35), CTCL versus AD (n = 37), and MPR versus AD (n = 39). Colored dots are significant DEPs, dashed vertical lines indicate log2 fold change of ≤ 0.5 or ≥ 0.5. Numbers indicate DEPs in each direction with log2 fold change of ≤ 0.5 or ≥ 0.5 in brackets. **d.** Overrepresentation analysis (Reactome database) of t-test significant proteins in psoriasis versus PRP (log2 fold change ≤ 0.5 or ≥ 0.5) (n = 35). **g.** Number of ANOVA / post-hoc Tukey HSD significant proteins per comparison. n = 93 individuals. **h.** Selected ANOVA significant proteins in AD versus CTCL (n = 37) and PRP versus psoriasis (n=35). **a, h.** Box plots show the median (center line) with interquartile range of 25% to 75%, whiskers extend to further data points. **c - h.** Benjamini-Hochberg correction for multiple comparisons (FDR < 0.05).

A second diagnostically challenging pair - cutaneous T-cell lymphoma (CTCL) and atopic dermatitis (AD) - also displayed distinct molecular profiles. CTCL showed increased expression of lymphocyte activation and survival markers (IL16, CXCL13, ZAP70, CD48, RASAL3) and enhanced interferon signaling (ISG20, STAT1) compared to AD (Fig. 2E). In contrast, AD was characterized by proteins associated with eosinophilic/Th2-driven inflammation (DUOX1, EPX, PSTN; Fig. 2E) and dominant IL-4/13 signaling (Extended Data Fig. 2L).

Our proteomic data also distinguished AD from MPR. MPR exhibited higher levels of interferon-responsive proteins (WARS1, GBP1/2, IFIT3, ISG15, STAT1), while AD showed upregulation of extracellular matrix protein COL6A6 and exonuclease TREX2. This is interesting because TREX2, selectively expressed in keratinocytes, is known to aggravate inflammation in psoriatic skin and small-molecule inhibitors are currently in development for inflammatory skin diseases (Fig. 2F) (*28–30*).

We next employed multi-condition analysis to identify disease-specific protein signatures with the highest specificity across the cohort (ANOVA, Fig. 2G, H; Extended Data Fig. 2J). This revealed elevated levels of keratinocyte alarmins (KRT6C), as well as the metabolic enzyme AKR1B10 and calcium-binding, visinin-like protein 1 (VSNL1) in psoriasis compared to PRP. Due to their role in cellular metabolism, AKR1B10 and VSNL1 overexpression may contribute to the characteristic hyperproliferative phenotype observed in psoriasis (*31, 32*). PRP expressed barrier protection proteins, including protease inhibitor SERPINB12 and antimicrobial peptide RNASE7 more abundantly (Fig. 2G, H), suggesting barrier dysfunction as a central and unexplored pathogenic mechanism in this disease.

In AD compared to CTCL, proteins associated with eosinophilic inflammation (EPX, POSTN) were again strongly enriched. Additionally, we detected higher expression of Charcot-Leyden crystal protein (CLC), formed from the eosinophil granule protein galectin-10 and strongly associated with type 2 immune responses (Fig. 2G, H). CLCs act as damage-associated molecular patterns (DAMPs) that activate the NLRP3 inflammasome, the latter contributing to chronic inflammation in AD (*33, 34*). In contrast, CTCL showed higher levels of PTPRCAP, a positive regulator of the lymphocyte surface marker CD45 (Fig. 2G) along with other proteins associated with lymphocyte activation and survival (IL16, CXCL13, CD48, ZAP70; Fig. 2H). Notably, the secretion of IL16 by malignant T-cells is associated with disease progression, and therefore offers diagnostic potential (*35, 36*).

### Transcriptome of erythrodermic skin reveals disease-specific patterns

Having established disease-specific protein signatures, we next analyzed the transcriptome of the cohort. We detected ∼62,000 unique transcripts across all samples, a quarter of which were protein-coding (27.4%, n = 17,255; Extended Data Fig. 4A-D). PCA separated healthy controls from disease samples along PC1 (12.8% variance), while PC2 (8.5% variance) segregated disease entities similar to our proteomics findings (psoriasis/PRP vs. CTCL/MPR; Extended Data Fig. 4E). Interestingly, transcripts involved in epidermal differentiation and barrier function, such as suprabasin (SBSN) and ceramide synthase 3 (CERS3), were the main drivers separating between diseases and along PC2 (Extended Data Fig. 4F).

Pairwise comparison identified numerous differentially regulated transcripts (DETs; Extended Data Fig. 4G, H). Pearson correlation of DETs across all diseases revealed specific clusters for PRP, psoriasis, CTCL and MPR (Fig. 3A). The PRP-associated cluster (#2), contains a prominent interleukin-17C (IL17C) and CC chemokine ligand-20 (CCL20) signature compared to other diseases (Fig. 3B, C). This was unique to IL17C, while other IL17-family members did not have a PRP-specific expression pattern (Extended Data Fig. 4I). Interestingly, we also observed heterogeneity among PRP patients based on their IL17C expression levels, suggesting disease endotypes. Indeed, IL17C expression was strongly associated with multiple interleukin-1 family cytokines, including IL36B, IL1F10, IL1A, IL1B and IL1FN (Fig. 3D). In contrast, patients with lower IL17C expression tended to have higher levels of innate pattern recognition receptors (TLR3), TNF family members (TNFSF13B), and calcium-binding proteins (S100A6) (Fig. 3D).

**Fig. 3.**
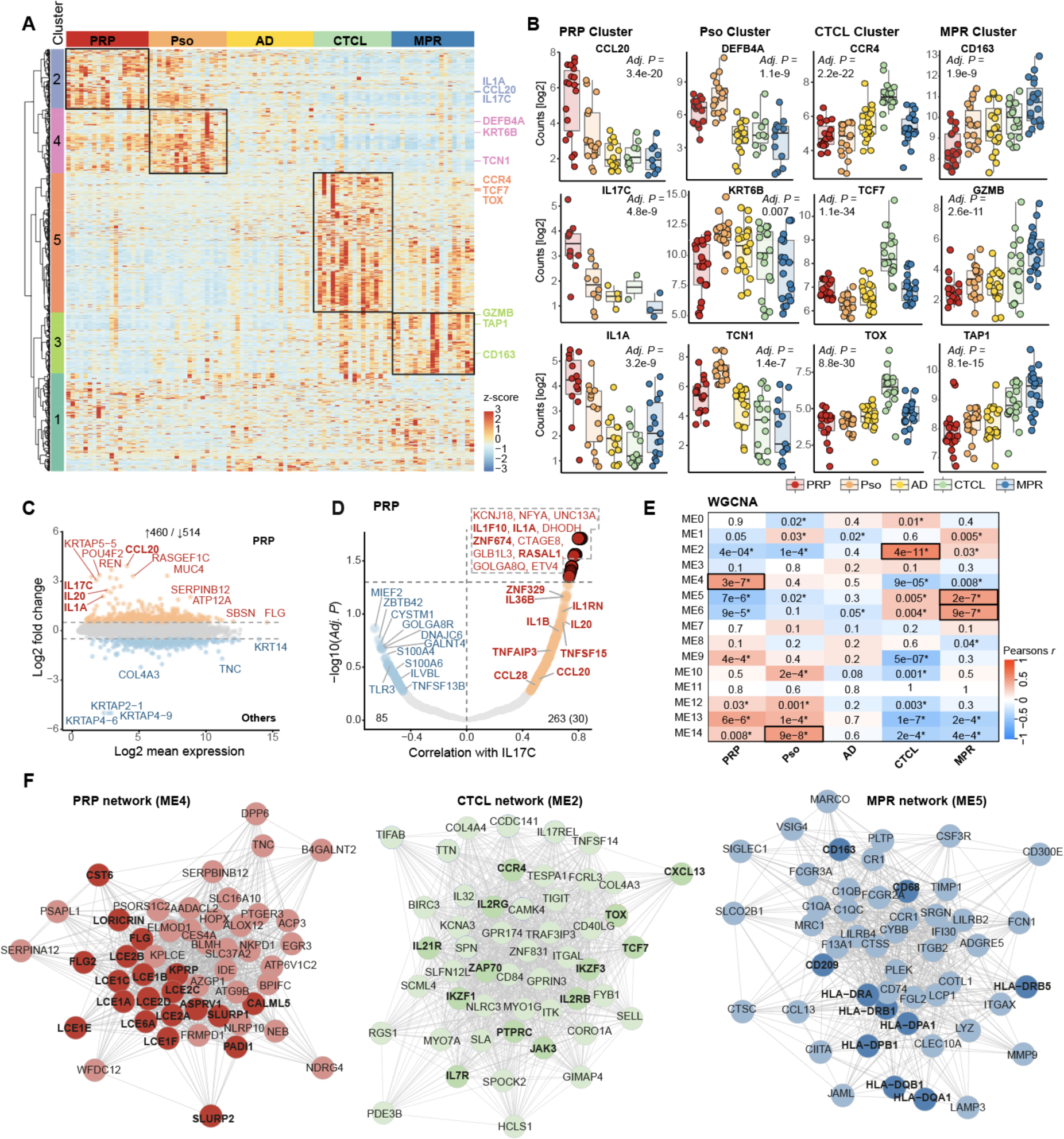
The transcriptome of erythrodermic skin diseases. **a.** Semi-supervised heatmap of t-test significant transcripts with log2 fold change ≤ 1.5 or ≥ 1.5. Color represents normalized intensity levels (z-score). **b.** Box plots of t-test significant transcripts from the different clusters in c., showing the median (center line) with interquartile range of 25% to 75%, whiskers extend to further data points. Significance is calculated by Likelihood Ratio (LRT) (n = 93 individuals). **c.** MA plot of differentially expressed transcripts (DETs) between PRP and the other diseases with log2 fold change ≤ 0.5 or ≥ 0.5. Numbers indicate DETs in each direction (n = 93 individuals). **d.** Transcripts correlating (Pearson r) with IL17C expression in PRP samples. Light colors indicate nominal significance (P < 0.05), and dark colors indicate significance after multiple testing correction (FDR < 0.05) (n = 19 individuals). **e.** Weighted gene co-expression analysis (WGCNA). Colors represent module correlations (Pearson’s r) and the numbers represent the corresponding Adj. P with * indicating significance (p <0.05) (n = 93 individuals). **f.** Network analysis of highly correlating modules with either PRP, CTCL or MPR. Transcripts of interest are highlighted in bold. **b-e** Benjamini-Hochberg correction for multiple comparisons (FDR < 0.05).

The CTCL-associated cluster (#5) revealed upregulation of CCR4, a chemokine receptor expressed on malignant T-cells and the target of mogamulizumab, an approved therapeutic monoclonal antibody in this disease (*37*). Among highly upregulated transcription factors we found TCF7 (Transcription Factor 7) and TOX (Thymocyte selection-associated high mobility group box protein), both implicated in the pathogenesis and prognosis of CTCL (Fig. 3A, B; Extended Data Fig. 4H) (*38, 39*).

The psoriasis-associated cluster (#4) was characterized by overexpression of antimicrobial peptides (DEFB5A), alarmins (KRT6B) and TCN1, corroborating our proteomics findings (Fig. 3A, B). Notably, lipocalin 2 (LCN2) and mucin-4 (MUC4) were among the most highly upregulated transcripts in psoriasis compared to all other diseases (Extended Data Fig. 4H). While LCN2, a mediator of neutrophil chemotaxis, has previously been implicated in psoriasis (*38*), our data also suggests an association between MUC4 and psoriasis. MUC4 regulates the expression of LCN2, and this axis may contribute to the neutrophilic infiltration in psoriatic lesions (*40*). Lastly, the MPR-associated cluster (#3) demonstrated upregulation of interferon-induced chemokines (CXCL9-11), MHC class molecules (HLA-B), antigen presentation transporters (TAP1) and cytotoxic effector molecules (GZMB, GNLY), all supportive of an active T-cell-mediated immune response known to be involved in drug hypersensitivity reactions (Fig. 3A, B; Extended Data Fig. 4H).

To explore coordinated transcriptional programs, we performed weighted gene co-expression analysis (WGCNA, (*23*)) and identified distinctive networks for each disease (Fig. 3E). The psoriasis network predominantly featured transcripts associated with epidermal hyperproliferation (KRT6A-C, KRT16, KRT17, IL36G) and antimicrobial defense (DEFB4A, S100A7-9, PI3). In contrast, the PRP network was enriched for skin barrier proteins (LCE family, filaggrin, loricrin, CALM5, CST6, SLURP1), suggesting altered epidermal differentiation and barrier dysfunction as a contributing pathogenic mechanism. Notably, several of these transcripts are involved in sphingolipid metabolism, a pathway previously implicated in psoriasis but not explored in PRP (Fig. 3F; Extended Data Fig. 5A) (*41*). The CTCL network exhibited pronounced T cell proliferation and activation, including the previously identified CCR4, TOX, TCF7 and CXCL13, further highlighting their importance in CTCL pathogenesis (*37, 38, 42, 43*) (Fig. 3F; Extended Data Fig. 5C). For MPR, we identified two distinct networks, including one primarily associated with monocyte signaling and antigen presentation, and another with interferon signaling and viral response mechanisms (Fig. 3F; Extended Data Fig. 5B, D).

### Multi-omics analysis reveals shared disease signatures

To uncover molecular signatures consistently altered in both omics modalities, we first performed direct integration of matched transcript-protein pairs. We observed an overlap of > 6,000 pairs, representing a third of all protein-coding transcripts and the majority of proteins (Fig. 4A). Consistent with previous multi-omics studies (*44*), transcript counts and protein intensity levels showed only a low degree of correlation (Pearson’s *r* = 0.3) (Fig. 4A, B). In contrast, disease-specific biological alterations (i.e., fold changes) were highly correlative between both modalities (Pearson’s *r* = 0.71 – 0.76), with up to 82,3% showing concordant regulation (Fig. 4C, D).

**Fig. 4.**
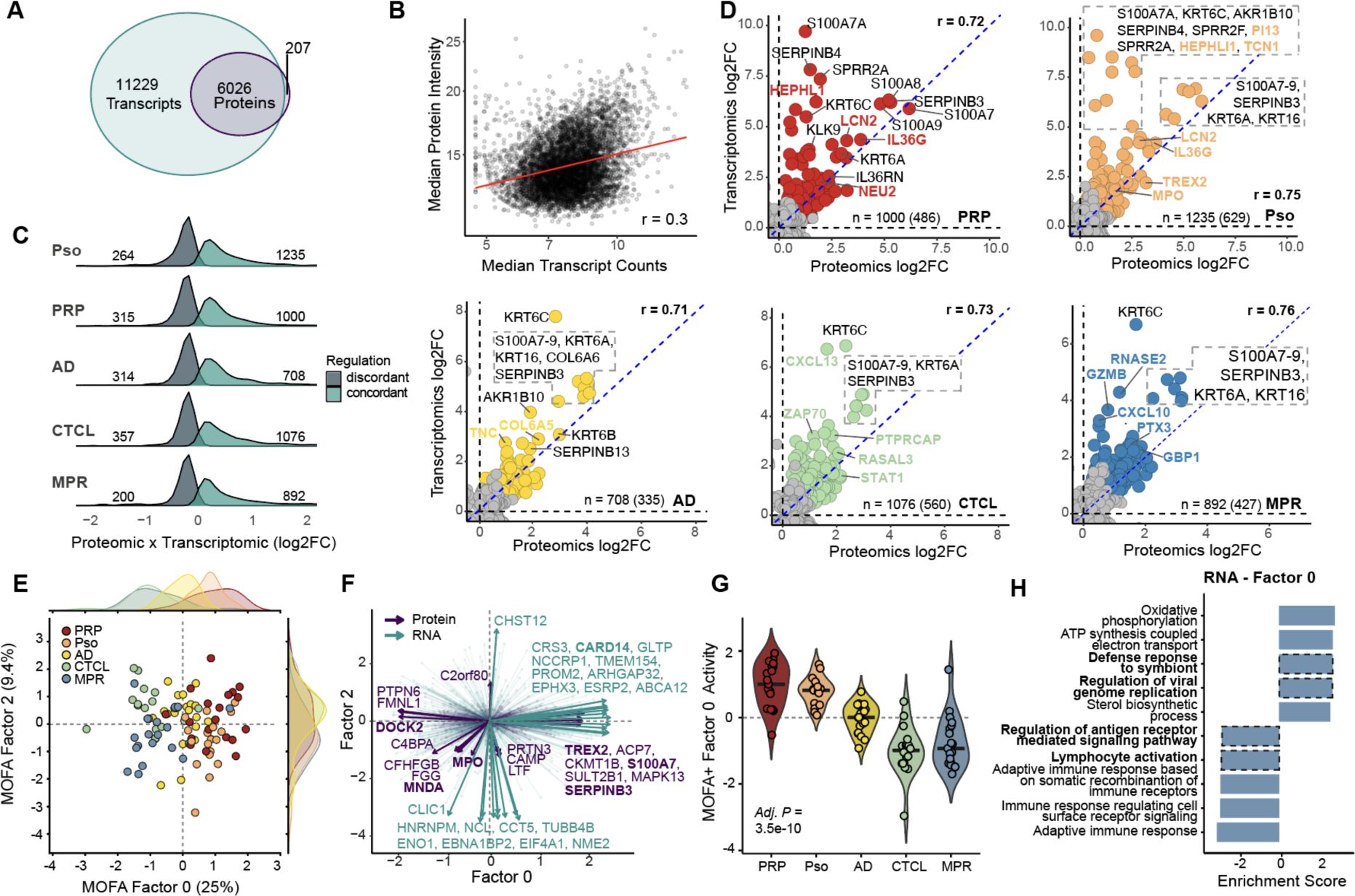
Multi-omics data integration. **a.** Number of overlapping transcripts and proteins. **b**. Scatter plot of median protein intensities and transcript counts. Pearson correlation (r) calculated based on intra-individual intensities. **c.** Ridge density plot showing the correlation between proteomics and transcriptomics log2 fold change of the different diseases versus healthy. Colors represent concordant or discordant regulation. **d.** Intersection of t-test significant proteins/ transcripts of each disease versus healthy. Different colors represent the cohort. Numbers indicate transcripts/proteins with positive correlation with log2 fold change of ≤ 0.5 or ≥ 0.5 in brackets. **e.** Dimensions of the MOFA interfered latent space for factors 0 and 2. Dots indicate patient samples and are colored based on the cohort. Distributions indicate the kernel density estimate of the sample distribution per cohort. **f.** Biplot of MOFA factors 0 and 2 with lines indicating direction and magnitude of contribution and the color the origin. **g.** Cohort-wise factor activity for MOFA factor 0. Significance is calculated by Kruskal-Wallis H test with Benjamin-Hochberg correction. **h.** Gene set enrichment analysis (GSEA) of transcript in factor 0 (GO Biological process). **g,h.** Benjamini-Hochberg correction for multiple comparisons (FDR < 0.05). **j,k.** Benjamini-Hochberg correction for multiple comparisons (FDR < 0.05). **a-h.** n = 91 individuals.

Across all type of erythroderma, numerous alarmins and defensins (S100A7-9, SERPINB3, KRT6A and KRT16) were upregulated at both the protein and transcript levels compared to healthy controls (Fig. 4D). In psoriasis, neutrophil markers (MPO and TCN1) were consistently elevated, while elevated barrier protein COL6A6 and type 2 immunity-associated TNC expression were dominant in AD (Fig. 4D) (*45*). In CTCL, shared markers included ZAP70, STAT1, RASAL3 and the chemokine CXCL13. Although RASAL3 has been implicated in cancer biology and is known to be expressed in T-lymphocytes, negatively regulating RAS signaling, its function in CTCL remains unexplored (*46*). In MPR, interferon-mediated response molecules (CXCL10, GBP1) and cytotoxic effector Granzyme B (GZMB) were robustly upregulated (Fig. 4D).

In multiple instances proteins were upregulated despite the level of the corresponding transcripts being decreased (Fig. 4C; Extended Data Fig. 6C, D). Notably, these discordantly expressed proteins and transcripts included a large number of ribosomal proteins (Extended Data Fig. 6B), suggesting specific post-translational regulatory mechanisms affecting protein synthesis machinery in inflammatory skin conditions such as erythroderma and its underlying causal diseases. Among these discordantly regulated molecules, three inflammation-associated proteins (TIMP3, HTRA1, DNAI4) were present in all cases, linking the TNF-α and NF-κB signaling pathways as common denominators of erythroderma and the underlying skin conditions PRP, psoriasis, AD, CTCL and MPR (Extended Data Fig. 6A) (*47, 48*).

Moving beyond one-to-one relationships, we applied multi-omics factor analysis (MOFA) to capture multivariate patterns across both datasets (*21*). Among the first seven factors, five were significantly associated with the underlying disease, and both the proteome and transcriptome contributed to these disease-related patterns (Extended Data Fig. 7A-D). The first factor (factor 0), driven two-thirds by the transcriptome and one-third by the proteome, primarily separated CTCL and MPR samples from PRP/psoriasis samples (Fig. 4E-G). Notably, the separation towards PRP/psoriasis was driven by the exonuclease TREX2 and CARD14, previously implicated in their respective pathogenesis (Fig. 4F) (*28, 49, 50*). In contrast, myeloid cell nuclear differentiation marker (MNDA) was a driver towards MPR, highlighting the importance of the innate immune system in this disease (*51*). The PRP/psoriasis cluster was further associated with defense responses to symbionts and regulation of viral genome replication, whereas CTCL/MPR was associated with antigen receptor signaling and lymphocyte activation (Fig. 4H; Extended Data Fig. 7G). Subsequent MOFA factors two, three, five and seven provided more nuanced discrimination patterns (Fig. 4E, G; Extended Data Fig. E, F, H). One such example is factor five, revealing DUOX1 and TNC as main drivers of AD separation (Extended Data Fig.7H).

### Machine learning based diagnostic platform for disease stratification in erythroderma

Classification of erythroderma for optimal therapeutic decision-making is a significant diagnostic challenge due to the frequent absence of clinical or histopathological signs specific for the causal underlying diseases. Consistent with the published literature (*4*), blinded histopathological evaluation of our cohort by board-certified dermatopathologists achieved diagnostic accuracy of 59.5% whereas clinical diagnosis alone was only achieved in 56.8% of cases (Extended Data Fig. 1B, C). To investigate whether our multi-omics data could enhance diagnostic accuracy in erythroderma, we extended our recently developed ADAPT-MS (Adaptive Diagnostic Architecture for Personalized Testing by Mass Spectrometry) pipeline to handle multi-modal data, now termed ADAPT-Mx (Fig. 1) (*24*). This approach dynamically retrains classification models for each individual sample, overcoming fundamental limitations in translating discovery-type multiomic measurements into relevant clinical decision-making.

To this end, we stratified the patient cohort into a balanced discovery (n = 48) and validation cohort (n = 47) and re-processed the MS-Raw data separately for those to ensure robust model development and independent testing without data leackage.

From the discovery cohort, we performed feature selection by selecting the top n significant proteins for t-tests between each of the disease groups with n=200 as an optimal value. Feature selection was performed using 10x bootstrapping within five cross-validation splits, to capture cohort and sample heterogeneity. This yielded a preselected set of 1,744 discriminatory features, including 1,068 transcripts (2% of all transcripts) and 676 proteins (7% of all proteins), with excellent classification performance on the discovery cohort (Fig. 5A). Notably, important molecular drivers of disease classification were related to intercellular communication and skin architecture, including COL6A6, CXCL10, CXCL11 and IL1A, amongst others. We then evaluated the predictive performance of our model in the independent validation cohort using ADAPT-Mx, by fitting the logistic regression classifier for each sample based on the available features, without imputation. It maintained its strong discriminatory power, with AUC values between 0.87 and 0.99 across all disease entities, thus showing a high degree of generalization of the model (Fig. 5B; Extended Data Fig. 8). Precision-recall analysis, which measures the accuracy of positive predictions for each specific disease, confirmed the robust performance of our model, with average precision scores ranging from 0.76 to 0.98 (Fig. 5C). From the broad list of selected features in the validation procedure, we found that 945 features (54%) were used for every single sample classification (Fig. 5D).

**Fig. 5.**
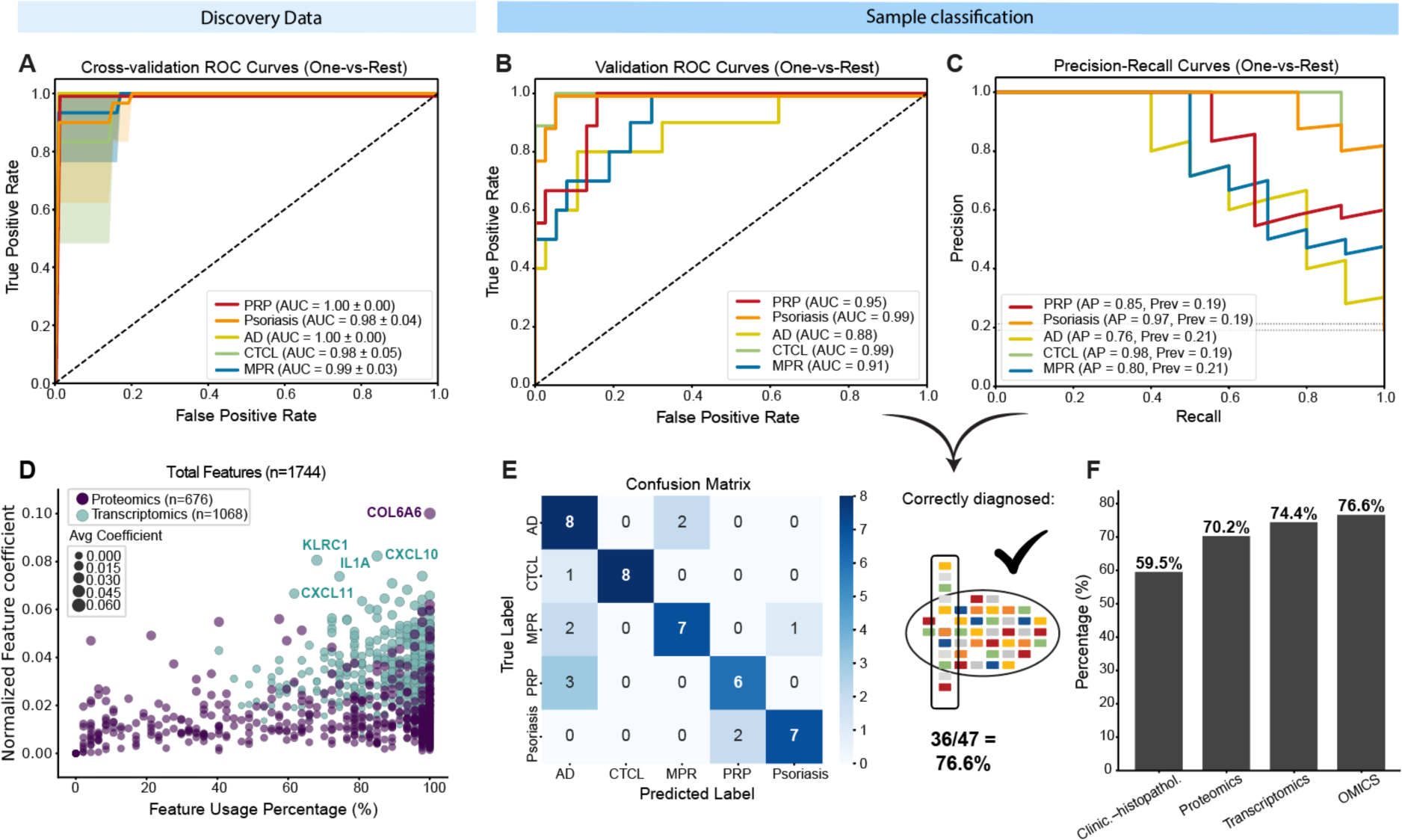
ADAPT-Mx Classification of Erythrodermic Skin Disease. **a.** Model performance for the discovery cohort (n = 48) shown as receiver operating characteristic (ROC) curves with area under the curve (AUC) values for each disease. **b,c.** Model performance for the validation cohort (n = 47) assessed through **(b)** ROC curves and AUC values and **(c)** disease-specific diagnostic accuracy (AP, average precision). **d.** Feature importance vs usage for validation cohort. Dot color indicates the feature origin and dot size the average coefficient. **e.** Confusion Matrix showing the number of correctly diagnosed patients in the validation cohort (n = 47). **f.** Comparative diagnostic performance of machine-learning based classification based on proteomics, transcriptomics or combined proteomics and transcriptomics (‘OMICS‘) compared to conventional clinical-histopathological assessment. Pityriasis rubra pilaris (PRP), psoriasis (Pso), atopic dermatitis (AD), cutaneous T-cell lymphoma (CTCL) or maculopapular rash (MPR).

The overall accuracy of our model reached 76.6% when using the data from both proteomic and transcriptomic modalities (Fig. 5E), slightly exceeding each single-modality approach, while clearly outperforming the accuracy of the current gold-standard clinical-pathological correlation (Fig. 5F, Extended Data Fig. 1B, C).

## Discussion

Timely and accurate diagnosis of the underlying inflammatory or malignant skin disease causing generalized skin redness and scaling (erythroderma) represents a profound challenge in dermatology. Given the severe impact on quality of life caused by a myriad of symptoms, including itch, fever, tachycardia, hypotension and increased risk of infection with potential for fatal outcome, new diagnostic modalities are needed to enable rapid targeted therapeutic intervention. To date, the gold-standard for diagnosis remains clinical-histopathological evaluation, which is challenging due to the extreme similarity of the underlying diseases when presented as erythroderma at both clinical and histological levels. Here, we applied in-depth multi-omic profiling of 116 FFPE patient biopsies of erythrodermic skin, caused by pityriasis rubra pilaris, psoriasis, atopic dermatitis, cutaneous T-cell lymphoma, or maculopapular drug rash, alongside healthy controls. Using MS-based proteomics and transcriptomics covering more than 17,200 transcripts and 9,300 proteins, we identified biological mechanisms and defined molecular signatures that improve speed and accuracy of diagnosis and hold potential for targeted therapy.

At a diagnostic level, a key innovation of this study is a novel molecular-driven diagnostic workflow - ADAPT-Mx - that uses our patient biopsy-derived multimodal omics data to dynamically retrain classification models for each individual, enabling improved clinical decision-making. This approach provides a robust foundation for clinical application and supports adoption at other healthcare institutions. With AUC values between 0.87-0.99 across all disease entities, this molecular classifier substantially outperforms the diagnostic accuracy of the current gold-standard clinical-histopathological correlation. While integrating both proteomics and transcriptomics achieved the highest accuracy in our validation cohort (76.6%), the model maintained strong performance even when using a single modality, facilitating its implementation based on the available resources. With additional validation in larger cohorts, this approach could fundamentally change how erythrodermic conditions are diagnosed, leading to more rapid, accurate and effective patient care.

The power of our combined omics analysis of skin biopsies also uncovered disease-specific pathways and molecular targets with potential therapeutic relevance. For instance, the IL17C-CCL20 axis was highly and selectively upregulated in erythrodermic pityriasis rubra pilaris. While high levels of IL17C have previously been shown in inflamed skin (*52, 53*), our data demonstrates its specificity for pityriasis rubra pilaris even among other severe inflammatory skin diseases. Furthermore, other members of the IL17 family were not elevated in pityriasis rubra pilaris. IL17C is produced by keratinocytes and signals through the IL-17RA/RE receptor to trigger innate defense mechanisms, including CCL20 induction (*54*). We identified two pityriasis rubra pilaris endotypes based on IL17C expression levels, with IL17C^high^-patients showing strong correlation with IL1-family cytokines. These endotypes may explain the variable clinical response to therapeutic interventions using IL1 and IL17 inhibitors (*27*) and suggest that molecular stratification could improve treatment selection. These results strengthen the growing evidence that agents targeting IL17C signaling (e.g., brodalumab, which blocks the IL-17RA receptor) may be more effective than classical IL17A inhibitors for certain pityriasis rubra pilaris patients.

Both transcriptomic and proteomic analyses independently linked erythrodermic pityriasis rubra pilaris to altered barrier function and aberrant keratinocyte differentiation. Network analysis of our transcriptomic data revealed pronounced dysregulation of sphingolipid metabolism pathways, while proteomic profiling showed significant upregulation of KRT23, a stress-inducible keratin involved in cellular proliferation and DNA damage response (*55–57*). While abnormal sphingolipid metabolism has been implicated in the pathogenesis of psoriasis, its role in pityriasis rubra pilaris remains unexplored.

In erythrodermic atopic dermatitis, we identified elevated expression of CLC (Charcot-Leyden crystal protein), which results from the eosinophil granule protein galectin-10 and is strongly associated to type 2 immune responses (*33*). Notably, CLC can form crystals in tissues with eosinophilic inflammation (*58*), and has been shown to activate the NLRP3 inflammasome in other allergic conditions (*33*), though this specific mechanism has not been directly demonstrated in AD. Given that NLRP3 plays a role in AD pathogenesis through both inflammasome-dependent and independent pathways (*34*), this finding suggests a potential molecular link that warrants further investigation.

Precision medicine in dermatology is increasingly important given the biological diversity and clinical heterogeneity of skin diseases, which often require long-term management. Traditional “one-size-fits-all” approaches frequently fail as they cannot account for individual variation in disease mechanisms, treatment response, and comorbidities. In this study we provide a template for personalized medicine in dermatology and demonstrate this in severe and challenging cases of erythroderma. We envision that similar approaches can contribute to improve diagnostic accuracy and identify molecular signatures for targeted therapeutic interventions across all skin diseases.

## Material and Methods

### Cohort Assembly

We retrospectively assembled a cohort of 116 formalin-fixed and paraffin embedded (FFPE) tissue sections from patients suffering from erythroderma (n = 96) and healthy controls (n = 20). The samples were obtained from the Ludwig-Maximilian-University (LMU) Munich (Germany), Johannes Wesling Medical Center Minden (Germany), University Hospital of Cologne (Germany) and histology center Kempf & Pfaltz (Zurich). All samples were re-evaluated histologically by a board-certified dermatopathologists (K.K.-F.) using a freshly cut and stained hematoxylin & eosin (H&E) section, and clinically through review of medical records and clinical images. For all patients, clinical metadata including age, sex and disease-specific scores (e.g. EASI for atopic dermatitis, PASI for psoriasis) were collected. In addition, the culprit drug (when identified) was documented for maculopapular rash (MPR), and disease stage (TNM) / blood involvement for cutaneous T cell lymphoma (CTCL). Healthy skin samples were either retrospectively or prospectively collected at the University Hospital of the LMU Munich. The study was ethically approved by the local ethics committee (22–0342, 22–0343).

### Clinical-Histopathologic Evaluation

To establish baseline diagnostic accuracy, two board-certified dermatopathologists and experienced clinicians (R.S., K.K.-F.) independently evaluated clinical images and corresponding H&E-stained histopathological sections, in a blinded manner. Diagnostic accuracy was determined by comparing their diagnoses against the validated clinical diagnosis established through long-term patient follow-up (Extended Data Fig. 1B,C).

### Sample preparation for proteomic analysis

A 5 µm section from each FFPE block was cut onto a PEN membrane slide pre-treated with Vectabond (VectorLabs) and deparaffinized, as previously described (*59*). After air-drying, the tissue was excised along with the underlying membrane and transferred into individual wells of a 96-well plate, prefilled with 40 µl of lysis buffer (10% acetonitrile, 60 mM TEAB pH8.5, 5 mM TCEP, 25 mM CAA), leaving the last row empty. The plate was then sealed, centrifuged (500 × g, 1 min), heated (76°C, 30min), sonicated (Covaris LE220plus, per row: 300s, 450W peak and 225W average power, 50% duty factor, 200 cycles) and heated again (76°C, 30min). Then 200 ng of standard keratinocyte cell lysates (see below) was added to all empty wells of the last row. This was followed by overnight tryptic digestion using 0.5 ng of LysC/Trypsin per well, and quenched with TFA (final concentration 1%). Protein concentration was measured using the tryptophan assay (wavelength 350nm, Tecan plate reader). 200 ng of each sample was loaded onto Evotip Pure tips (Evosep) as per the manufactureŕs recommendation. All steps involving liquid dispensing were performed using a liquid handling robot (OT-2, Opentrons Labwork Inc).

### Standard keratinocyte lysate

Keratinocyte lysate (NHEK/SVTERT3-5) was prepared by Evercyte GmbH using our specified lysis buffer (10% acetonitrile, 60 mM TEAB pH 8.5, 5 mM TCEP, 25 mM CAA) to ensure consistency with our experimental samples and future reproducibility. Protein concentration was measured (tryptophan assay) and samples were aliquoted to 5 ng/ml.

### LC-MS and raw data analysis

All samples were analyzed on an Evosep One LC system (Evosep Biosystems) coupled to an Orbitrap Astral mass spectrometer (Thermo Fisher Scientific). The standardized 60 SPD gradient (Evosep Biosystems) was applied on an 8cm PepSep column (150 μm diameter, 1.5 μm beads, # 1893470) heated to 50°C. The mass spectrometer was equipped with a FAIMS Pro interface (Thermo Fisher Scientific) operated at −40 V compensation voltage with 3.5 L min−1 total carrier gas flow, and EASY-Spray source (Thermo Fisher Scientific). Electrospray ionization was set to 2000 - 2400 V with RF Lens of 40%. MS1 acquisition covered 380 to 980 m/z with 240,000 resolution and 500% normalized automated gain control target. MS2 data were collected in narrow-window data-independent acquisition mode using 4 Th fixed windows with 5 ms maximum injection time. Fragmentation was performed by high-energy collisional dissociation at 25% normalized collision energy.

Raw data analysis was performed with DIA-NN v1.8.1 (*22*) searched against the human reviewed and TrEMBL FASTA database (taxonomy ID 9606), downloaded from Uniprot.org in February 2023. Variable modifications included are methionine oxidation and N-terminal acetylation, with cysteine carbamidomethylation applied as a fixed modification and a maximum of one missed cleavage. Precursor charge states were analyzed from 1 to 4, with m/z values spanning 300 to 1,800 and peptide lengths between 7 to 55 amino acids. Mass tolerances were set to 6 ppm for MS2 and 4 ppm for MS1. Match-between run (MBR) was enabled and all files were processed together, unless otherwise noted. Conservative protein grouping was achieved with the ‘--relaxed-prot-inf’ command. All other settings were as recommended by DIA-NN.

### Bulk RNA-seq

RNA was isolated from the FFPE tissue sections using the RecoverAll^TM^ Total NucleicAcid Isolation Kit (AM1975, Thermo Fisher Scientific, USA). Deparaffinization was performed using ROTICLEAR® (Carl Roth GmbH + Ko. KG, Germany) as per protocol. RNA concentration was measured using the Qubit^TM^ RNA High Sensitivity Assay (Thermofisher, USA). Sequencing libraries were created with 10 ng input per sample, using the SMARTer® Stranded Total RNA-Seq Kit v3 – Pico Input Mammalian (Takara Bio, Japan) and SMARTer® RNA Unique Dual Index Kit (Takara Bio, Japan), as well as NucleoMag DNA FFPE beats (Macherey-Nagel, Germany) per manufacturer’s protocol. Quality control was performed with the D5000 ScreenTape Assay (Agilent, USA) and libraries were sequenced with the NovaSeq 6000 (Illumina, USA) sequencing platform.

### Bioinformatics data analysis

FastQ files were processed on a high-performance Linux cluster using fastp for quality trimming and UMI extraction (*60*), Salmon for transcript quantification (*61*) and tximport for gene-level aggregation (*62*). Quality control was assessed with MultiQC (*63*). Subsequent bioinformatic analysis of both proteomic and transcriptomic data was performed within R (v 4.3.1) using RStudio.

For the transcriptomic data, only protein-coding transcripts with at least 3 counts in at least 50% of the samples per cohort were retained for subsequent analysis (before: 62,646; after: 17,255; Extended Data Fig. 4A). DESeq2 (v 1.44.0) was used for normalization and differential expression analysis (*64*). Normalization was performed with sequencing run as a covariate to account for potential batch effects. Outliers were identified and removed based on a threshold of 90% of the median (before: n = 115, after: n = 113). Differential expression analysis was conducted using the Likelihood Ratio Test (LRT), followed by log2 fold change shrinkage using the lfcShrink function with type “ashr” (*65*). Multiple testing correction was applied using the Benjamini-Hochberg (BH) method with a false discovery rate (FDR) cutoff of 1%. For visualization, variance stabilizing transformation (VST) was applied. Heatmaps were generated using the pheatmap package with zero-centered scaling. Weighted Gene Co-expression (WGCNA) analysis was performed on the top 50% variable transcripts (n = 110 individuals). Module detection was conducted with cutreeDynamic (deepSplit = 3, minClusterSize = 80) with a soft threshold of 13, targeting an adjusted R^2^ close to 0.9, followed by multiple testing correction (BH, FDR 5%) (*23*). Network analysis of the modules was performed with the igraph package (*66*).

For proteomic data, the pg.matrix.tsv output file from DIA-NN was filtered for >= 75% valid values within a cohort (before: 9382; after: 6233), followed by median imputation and batch correction using ComBat from the sva package (*67*). Outliers were removed based on median ±2 standard deviations in total protein intensity (before: n = 116; after: n = 113). Differential expression analysis was performed using limma (*68*) with measurement plate as a covariate, followed by multiple testing correction (BH, FDR 5%). Gene set enrichment analysis (GSEA) and over-representation analysis (ORA) were conducted with clusterProfiler (*69*) using the Reactome pathway database (filtered for: ‘interleukin’, ‘immune’, ‘cytokine’, ‘inflammatory’, ‘T cell’, ‘B cell’, ‘chemokine’, ‘interferon’) or the GO Biological Process database. For one-way analysis of variance (ANOVA), the R stats package was used, followed by multiple testing correction (BH, FDR 5%) and post-hoc Tukey Honest Significant Differences (Tukey-HSD) were calculated at a confidence level of 0.95. Correlations between both modalities were calculated using Pearson correlation based on either absolute or log2 fold changes (n = 110 individuals).

### Multimodal analysis with MOFA+

Multimodal analysis was performed in Python (version 3.12.8) with the muon (version 0.1.7, (*70*)) implementation of MOFA+ (version 0.7.2, (*21*)). Proteomics data were filtered, median-imputed, and batch-corrected as described before. The VST-transformed transcriptomics data was subset to match the number of features using the highly variable genes using the *scanpy.pp.highly_variable_genes* function (flavor=”seurat”, v.1.11.1). Both datasets were subsequently z-scaled (n = 91 individuals). MOFA+ was run on the union of all samples with Gaussian likelihood. The automatic relevance determination was enabled for both weights and factors (ard_weights=True, ard_factors=True), while the spike-and-slab prior was disabled (spikeslab_factors=False, spikeslab_weights=False). To determine the number of latent factors, the model was initially overfitted with *K* = 20 factors; it was then rerun with *K* = 12, corresponding to the number of factors explaining at least 3% of the total variance. To assess associations between latent factor activities and cohort-level covariates, we applied a non-parametric Kruskal–Wallis H test for categorical covariates (disease type, gender, location, plate/batch) and computed the Pearson correlation to continuous covariates (age). Resulting *P* values were corrected for multiple testing using the Benjamini–Hochberg procedure. Gene set enrichment analysis was performed on the factor weights with the decoupler package (v. 1.8.0, (*71*)) on the GO Biological Process pathway database.

### Adaptive diagnostic architecture for personalized testing (ADAPT)

For the application of the ADAPT ML framework, MS raw data were newly processed separately to avoid data leakage. The samples were randomly scrambled into a discovery set (50%; n = 48) and validation set (50%; n = 47) with equal distribution of all disease conditions prior to analysis. MS Raw files from the discovery set were processed using DIA-NN (v1.8.1) with the human protein database (SwissProtFASTA database; taxonomy ID 9606) at default settings in a two-step search including MBR (*22*). The spectral library created in the search of the discovery samples was then used to process all validation samples. For multi-omics classification, the transcriptomics features were appended to the proteomics data matrix either for all samples of the discovery set or within the single-sample processing pipeline of ADAPT on a per sample basis.

The ADAPT pipeline was performed in Python (version 3.12.8) using the scikit-learn library (version 1.1.3) for machine learning operations. The framework handles high-dimensional proteomics data with potential missing values through a systematic approach to feature selection and imputation. Prior to model development, we performed feature selection using pairwise statistical comparisons between all category combinations. For each pairwise comparison between cohorts, we conducted two-sample t-tests on the unimputed proteomics data, selecting features with significant differential expression. To account for multiple hypothesis testing, p-values were adjusted using the Benjamini-Hochberg false discovery rate (FDR) correction method. The top 200 features (proteins or transcripts) from each pairwise comparison were selected, creating a unified feature set that discriminates between all cohort combinations.

### Discovery cohort performance validation

We implemented a multiclass classification approach using logistic regression models. The model was trained using a stratified cross-validation framework to ensure balanced representation of all cohorts throughout the training process.

For each fold in the cross-validation, the following procedure was implemented: the data was split into 80% training and 20% test sets, maintaining the original class distribution. Feature selection was performed using only the training data to avoid data leakage. KNN imputation was applied to the training data. A logistic regression model was trained on the imputed data using only the selected features. The model was evaluated on the held-out validation data. This process was repeated for multiple runs (n = 3) with different random seeds to ensure stability of the results.

### Validation Framework

For validation samples the protein groups information of each sample was loaded from the single file search results, appended with the transcriptomics data and subjected to the ADAPT refitting architecture: In brief the overlap of selected features from the discovery data and the available features per sample is used to refit the logistic regression classifier on the discovery data set und subsequently classify the sample.

## Data Availability

All data produced in the present study are available upon reasonable request to the authors

## Acknowledgements

We thank all members of the Proteomics and Signal Transduction group at the Max Planck Institute of Biochemistry, especially Katharina Zettl for her support in the conduction of proteomic experiments, as well as Vincent Albrecht, Tim Heymann and Igor Paron for MS-method optimization and maintenance. Furthermore, RNA sequencing was performed by the following core facility: NGS Core Facility (RRID:SCR_025746). We thank the whole research group of Inflammatory Skin Diseases from the LMU Munich. We especially thank Claudia Kammerbauer for her support in performing the targeted transcriptomic experiments and Sabine Sirgez-Sasz for her support for collecting the FFPE blocks.

## Author contributions

L.E.F., P.-C.S., T.M.N. and Ma.M. conceived the project. P.-C.S. performed the proteomic and transcriptomic experiments together with T.M.N and M.Z.. P.-C.S. performed the statistical analysis of both datasets with help from T.M.N., L.D., G.W., M.N.-M., and J.B.M.-R.. L.D. performed the MOFA+ analysis. J.B.M.-R. performed the ADAPT analysis. P.-C.S., T.M.N., L.E.F. and Ma.M. designed the figures. P.-C.S., L.E., S.S., A.-S.B., and Mo.M. established the cohort and collected the clinical metadata. D.H., R.S., K.K.-F., W.K., Mi.M., A.L., B.K., V.G. and M.J.F. collected und validated the FFPE patient samples. P.-C.S., and N.M. collected the FFPE patient samples from the healthy controls. T.S., T.P., F.L., C.P., B.S., E.M.O., R.A.F., B.M.C.-E., S.C., S.F., N.J., I.K., J.L., A.S., A.O., Z.F., N. A.-Z., L.H.F., and M.W provided intellectual input and helped to interpret the data. P.-C.S., T.M.N., L.E.F. and Ma.M. wrote the manuscript. All authors read, revised and approved the manuscript.

---

**Extended Data Fig. 1.**
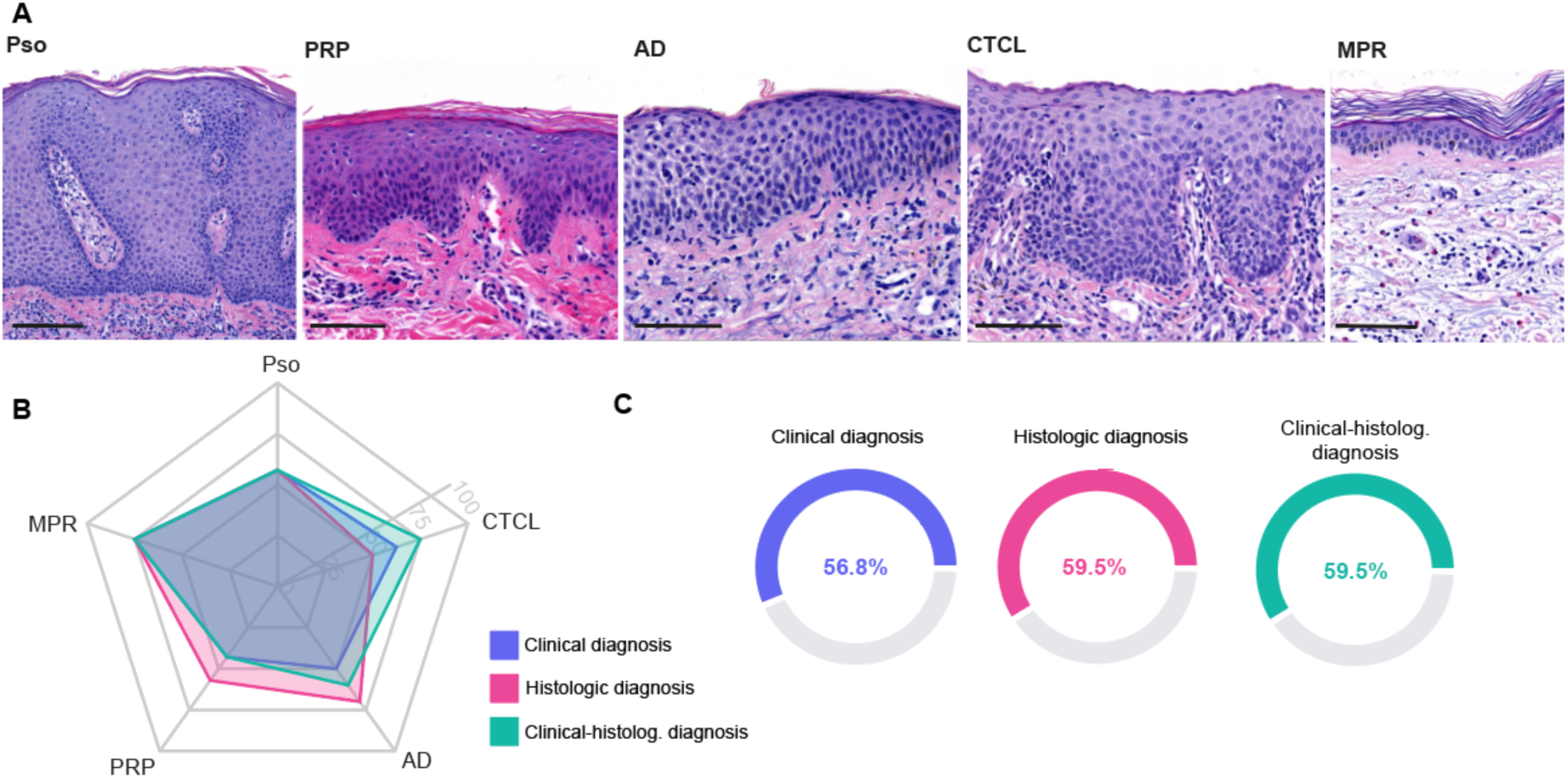
Clinical-histopathological correlation of erythrodermic skin diseases. **a.** Representative histological images of patients suffering from either psoriasis (Pso), pityriasis rubra pilaris (PRP), atopic dermatitis (AD), cutaneous T-cell lymphoma (CTCL), or maculopapular rash. Scale bar = 50 µm. b. Spider plot representing the percentage of correctly diagnosed patient cases after clinical and histological diagnosis, as well as clinical-histological correlation within the different diseases. Cases were analyzed by independent board certified dermato-histopathologists. c. Overall accuracy of the different methods for diagnosing erythroderma.

**Extended Data Fig. 2.**
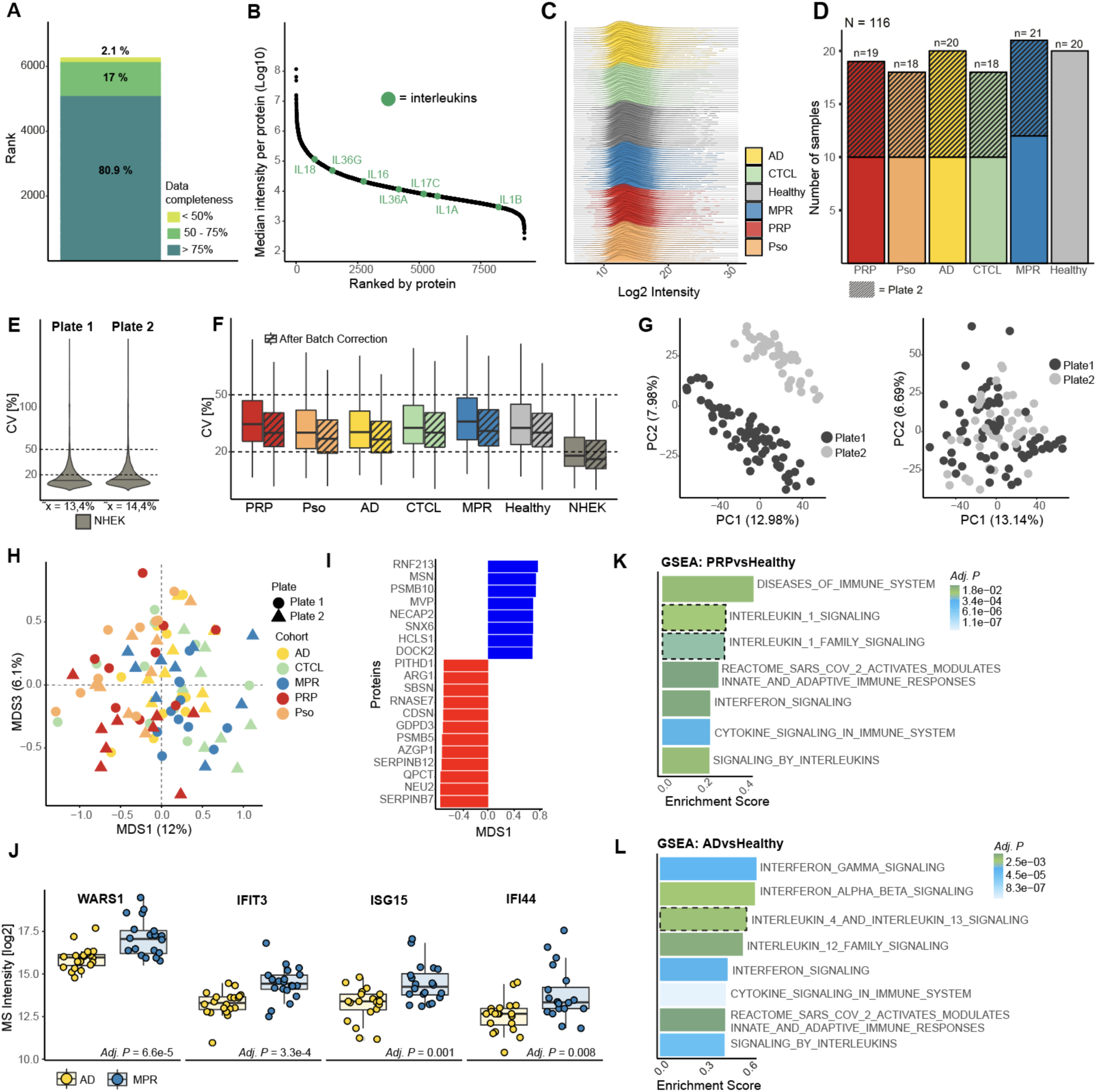
Analysis of proteomic data. **a.** Data completeness for each protein across all samples is represented by three groups (<50%, 50 – 75%, > 75%). n = 113 individuals. **b.** Abundance range rank plot of median protein intensity (y-axis). Interleukins are highlighted in green (n = 113 individuals). **c.** Intensity distribution (log2) across all samples or within individual samples (n = 113 individuals). **d.** Number of samples measured within plate 1 or plate 2 (marked with dashed lines). **e.** Median coefficient of variation (CV) of all proteins within the keratinocyte lysates (NHEK) within plates 1 and 2. n = 8 samples per plate. **f.** Median coefficient of variation (CV) of all cohorts between the plates (inter-plate variability) before and after batch correction. n = 113 individuals and 16 keratinocyte lysates (NHEK). **g.** Principal component analysis (PCA) of all patients before and after batch correction. Each dot is a patient, and colors represent the patients measured on the different plates (n = 113 individuals). **h.** Multidimensional scaling (MDS) of all patients. Each dot is a patient and color represent the corresponding cohort (n = 113 individuals). **i.** 20 top ranked loadings from MDS1. **j.** Box plots of ANOVA significant proteins in AD versus MPR. Box plots show the median (center line) with interquartile range of 25% to 75%, whiskers extend to further data points (n = 39 individuals). **k,l.** Gene set enrichment analysis (GSEA) of differentially expressed proteins in AD or PRP versus healthy (Reactome) (n = 38, upper panel and n = 39, lower panel). **j,k,l.** Benjamini-Hochberg correction for multiple comparisons (FDR < 0.05).

**Extended Data Fig. 3.**
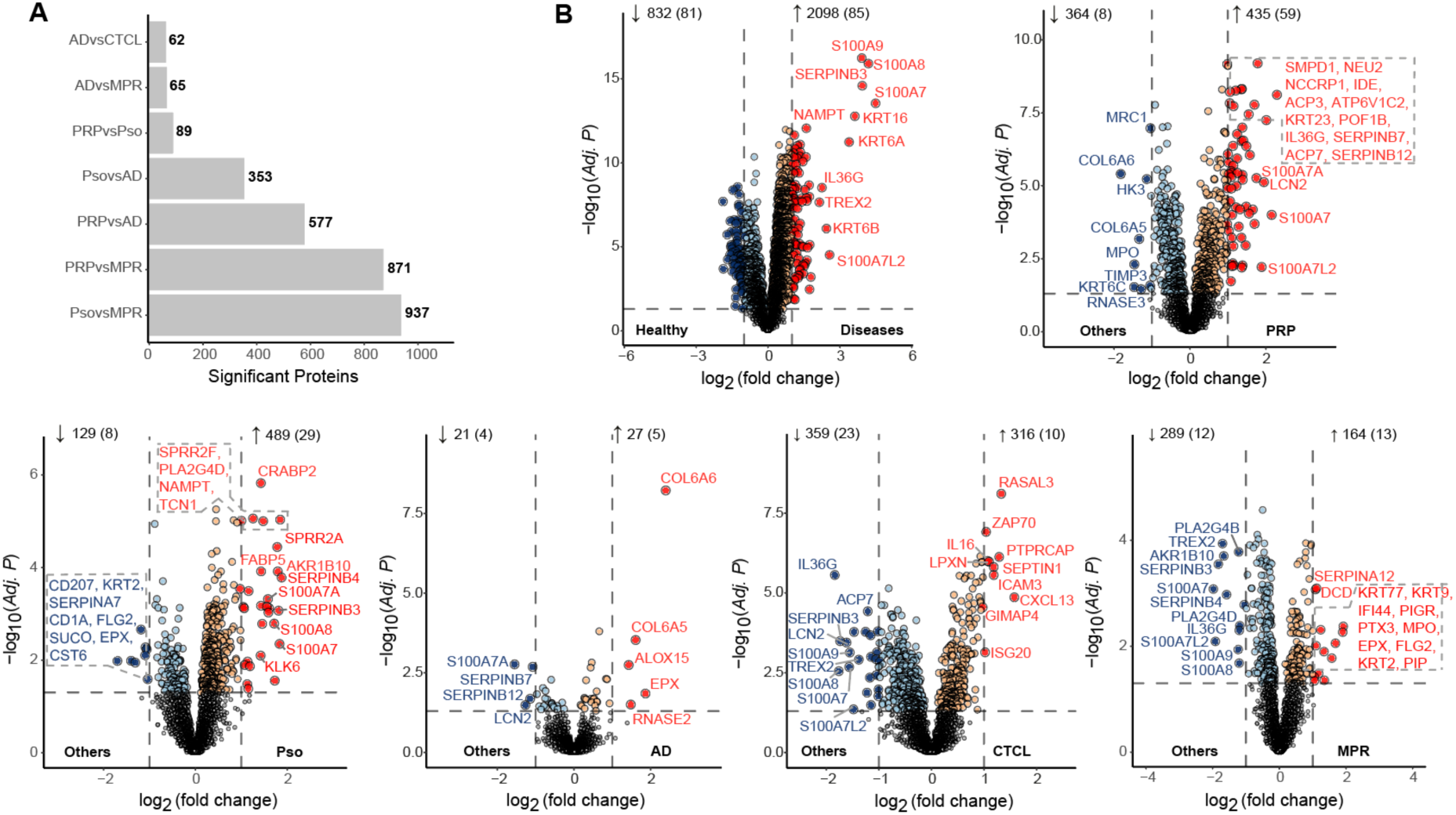
Differentially expressed proteins. **a.** Number of differentially expressed proteins (DEPs) within disease comparisons (n = 113 individuals). b. DEPs between all diseases versus healthy or a specific disease (PRP, Pso, AD, CTCL, MPR) versus all the others. Colored dots are significant DEPs, dashed vertical lines indicate log2 fold change of ≤ 1 or ≥ 1. Numbers indicate DEPs in each direction with log2 fold change of ≤ 1 or ≥ 1 in brackets. Benjamini-Hochberg correction for multiple comparisons (FDR < 0.05) (n = 113 individuals).

**Extended Data Fig. 4.**
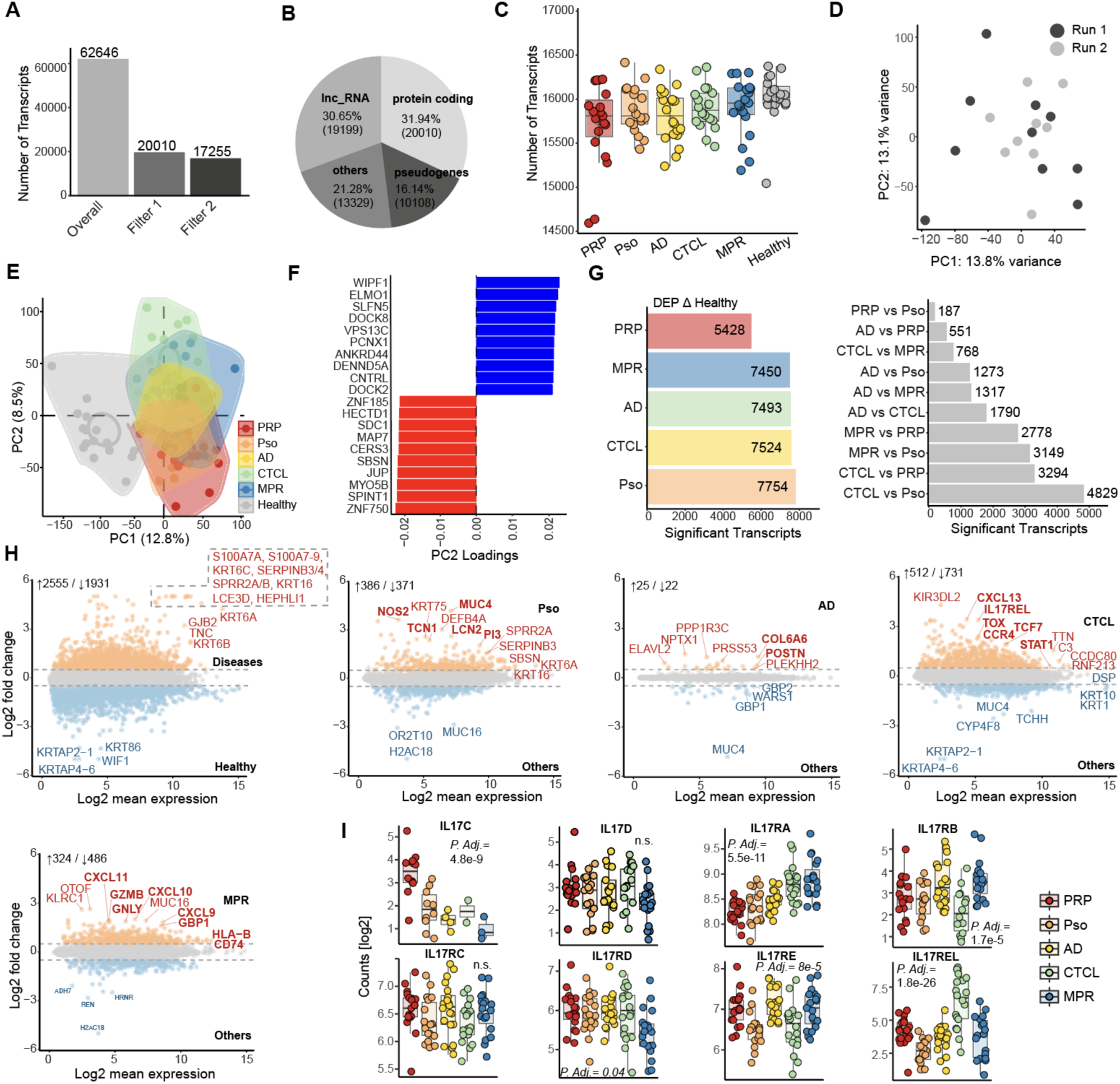
Analysis of transcriptomic data **a.** Number of measured transcripts overall, after filtering for protein-coding transcripts (filter 1) and after filtering for at least three counts in half of the individuals within a cohort (filter 2). **b.** Distribution of different types of measured transcripts. lnc_RNA, long non-coding RNA. **c.** Number of transcripts identified across the cohorts (n = 113 individuals). **d.** Principal component analysis (PCA) of pityriasis rubra pilaris (PRP) samples measured in two different runs. Each dot is a patient, and colors represent the patients measured on the different runs (n = 19 individuals). **e.** PCA of all patients (n = 113 individuals). Each dot is a patient, and color represents the corresponding cohort. **f.** Top 20 transcripts contributing to PC2. **g.** Number of differentially expressed transcripts (DETs) of the diseases versus healthy or within disease comparisons (n = 113 individuals). **h.** DETs between all diseases versus healthy or a specific disease (PRP, Pso, AD, CTCL, MPR) versus all the others. Colored dots are significant DETs, dashed vertical lines indicate log2 fold change of ≤ 1 or ≥ 1. Numbers indicate DETs in each direction with log2 fold change of ≤ 1 or ≥ 1 in brackets (n = 113 individuals). **i.** Box plots of IL-17 family members showing the median (center line) with interquartile range of 25% to 75%, whiskers extend to further data points. Significance is calculated by Likelihood Ratio Test (LRT) (n = 93 individuals).

**Extended Data Fig. 5.**
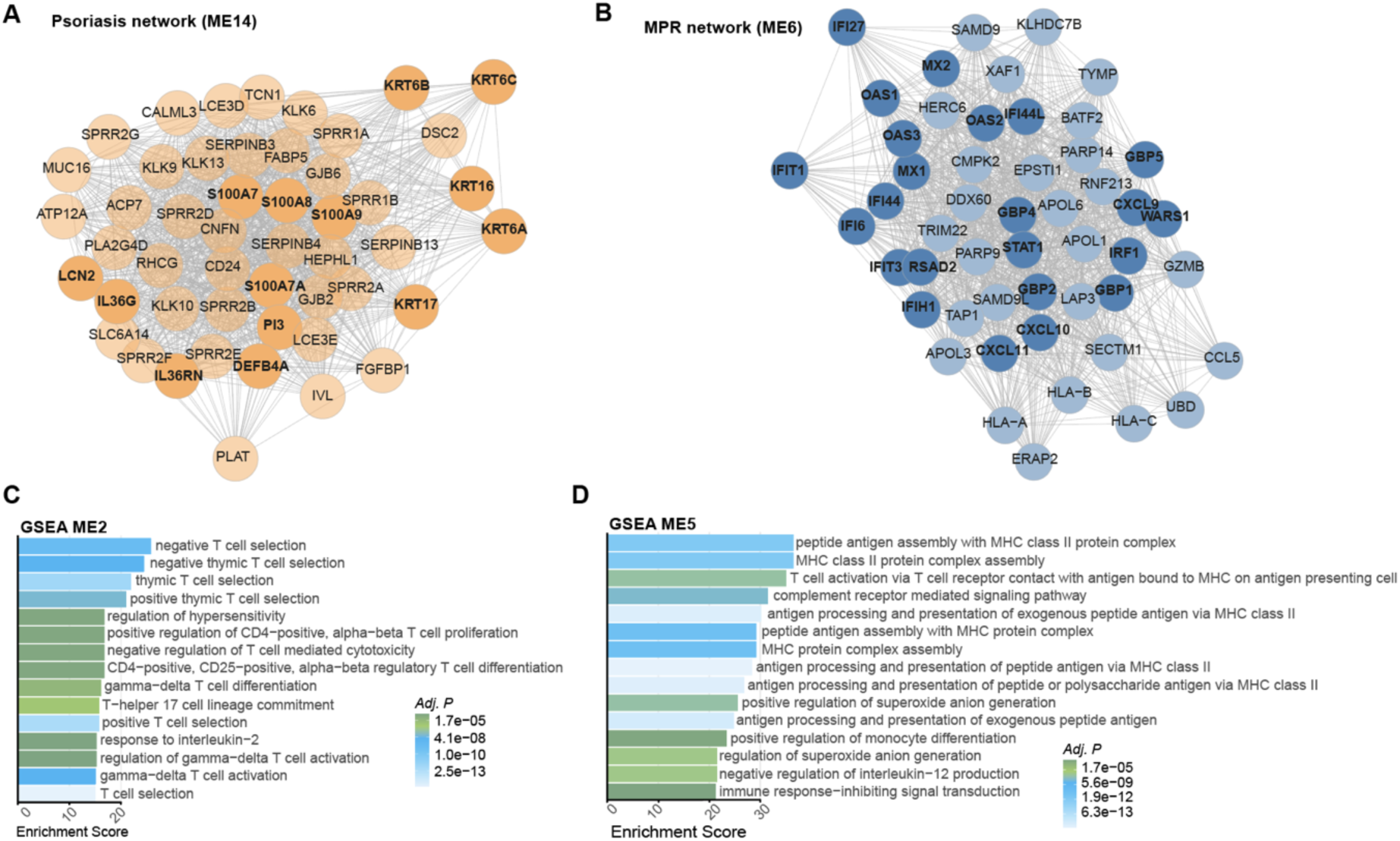
WGCNA analysis. **a-b.** Network analysis of highly correlating modules with either Pso or MPR. Transcripts of interest are highlighted by darker color and marked in bold. **c-d.** Gene set enrichment analysis (GSEA) of the CTCL module (ME2) and MPR module (ME5). Color represents significance level. **a-d.** Benjamini-Hochberg correction for multiple comparisons (FDR < 0.05) (n = 93 individuals).

**Extended Data Fig. 6.**
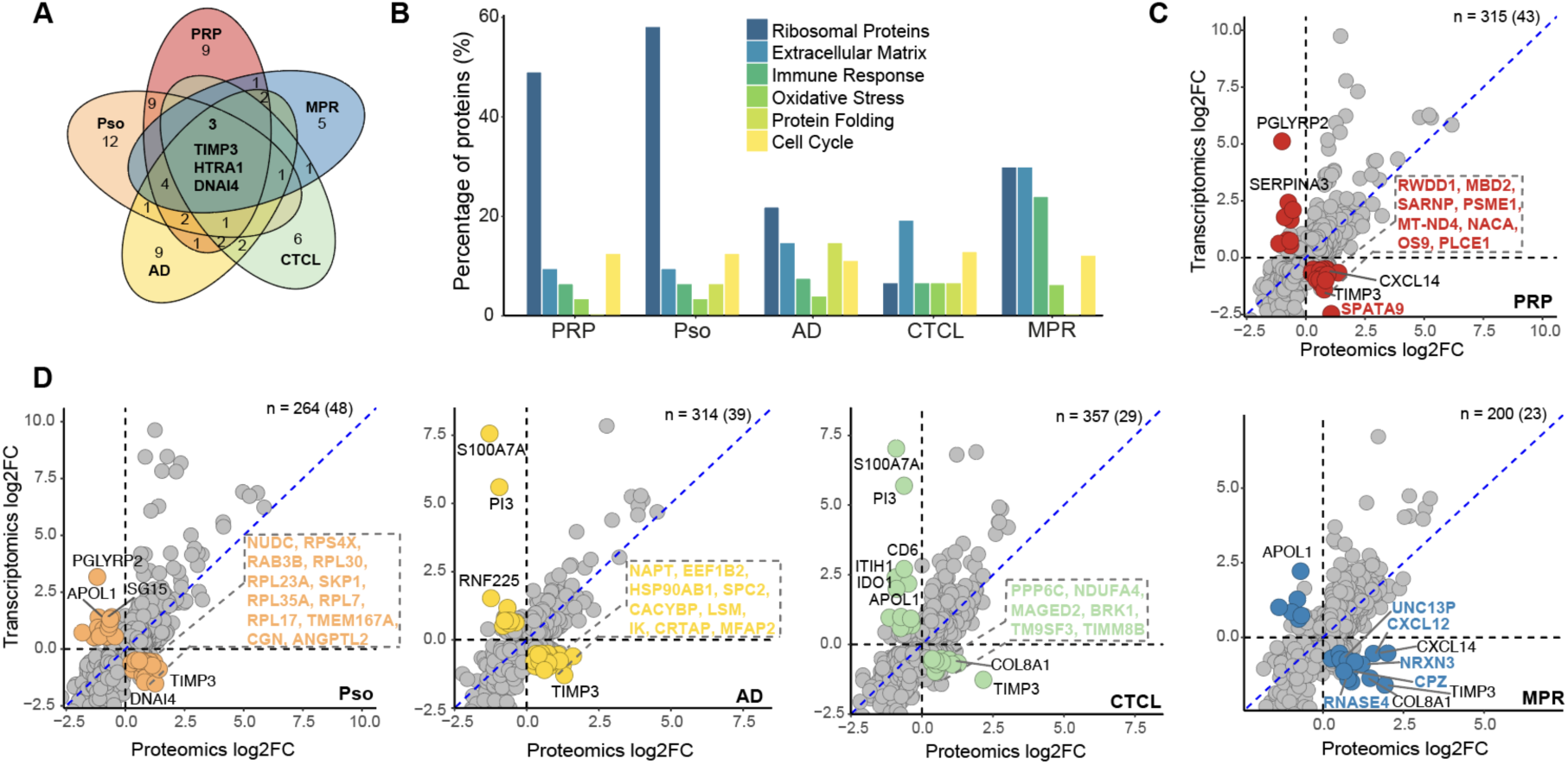
Omics data integration. **a.** Number of overlapping negatively correlated proteins (downregulated on transcriptional level) across different skin diseases. b. Barplot showing the distribution of negatively correlated proteins (downregulated on transcriptional level) across the different skin diseases, categorized by functional groups based on the GO biological process database. c-d. Intersection of t-test significant proteins/transcripts of each disease versus healthy. Different colors represent negative correlations within a specific cohort with log2 fold change of ≤ 0.5 or ≥ 0.5. Numbers indicate transcripts/proteins with negative correlation with log2 fold change of ≤ 0.5 or ≥ 0.5 in brackets (n = 91 individuals).

**Extended Data Fig. 7.**
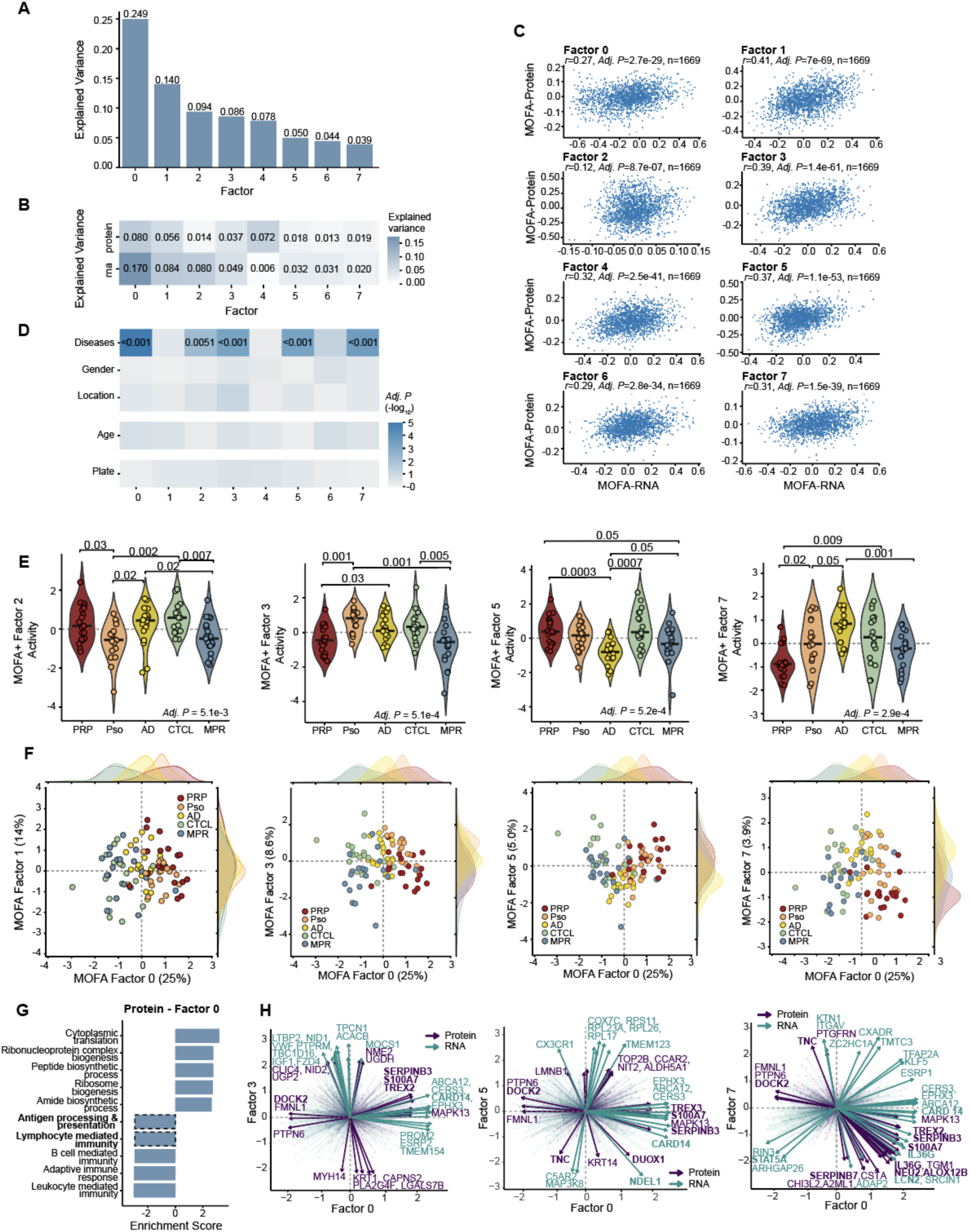
MOFA+ Analysis. **a.** Total explained variance per factor. **b.** Variance explained in each modality. **c**. Pearson correlation of MOFA+ weights between matched protein and RNA features (n = 1669) for each factor with corresponding significance value (Adj. P). **d.** Association of factor activity with cohort-level covariates. Annotations indicate factors with associations with Adj. P value < 0.05. Associations with categorical covariates (disease, gender, location, plate) were assessed using the non-parametric Kruskal–Wallis H test, while associations with continuous covariates (age) were evaluated using the t-statistic of the Pearson correlation coefficient. **e.** Cohort-wise factor activity for MOFA+ inferred factors 1, 3, 5 and 7. Significance is calculated by Kruskal-Wallis H test. **f.** Dimensions of the MOFA inferred latent space for factor 0, 1, 3, 5 and 7. Dots indicate patient samples and are colored based on the cohort. Distributions indicate the kernel density estimate of the sample distribution per cohort. **g.** Gene set enrichment analysis (GSEA) for the proteins in factor 0 (GO Biological process). **h.** Biplots of the different MOFA factors with lines indicating direction and magnitude of contribution and the color of the origin. **c-f.** Benjamini-Hochberg correction for multiple comparisons (FDR < 0.05) (n = 91 individuals).

**Extended Data Fig. 8.**
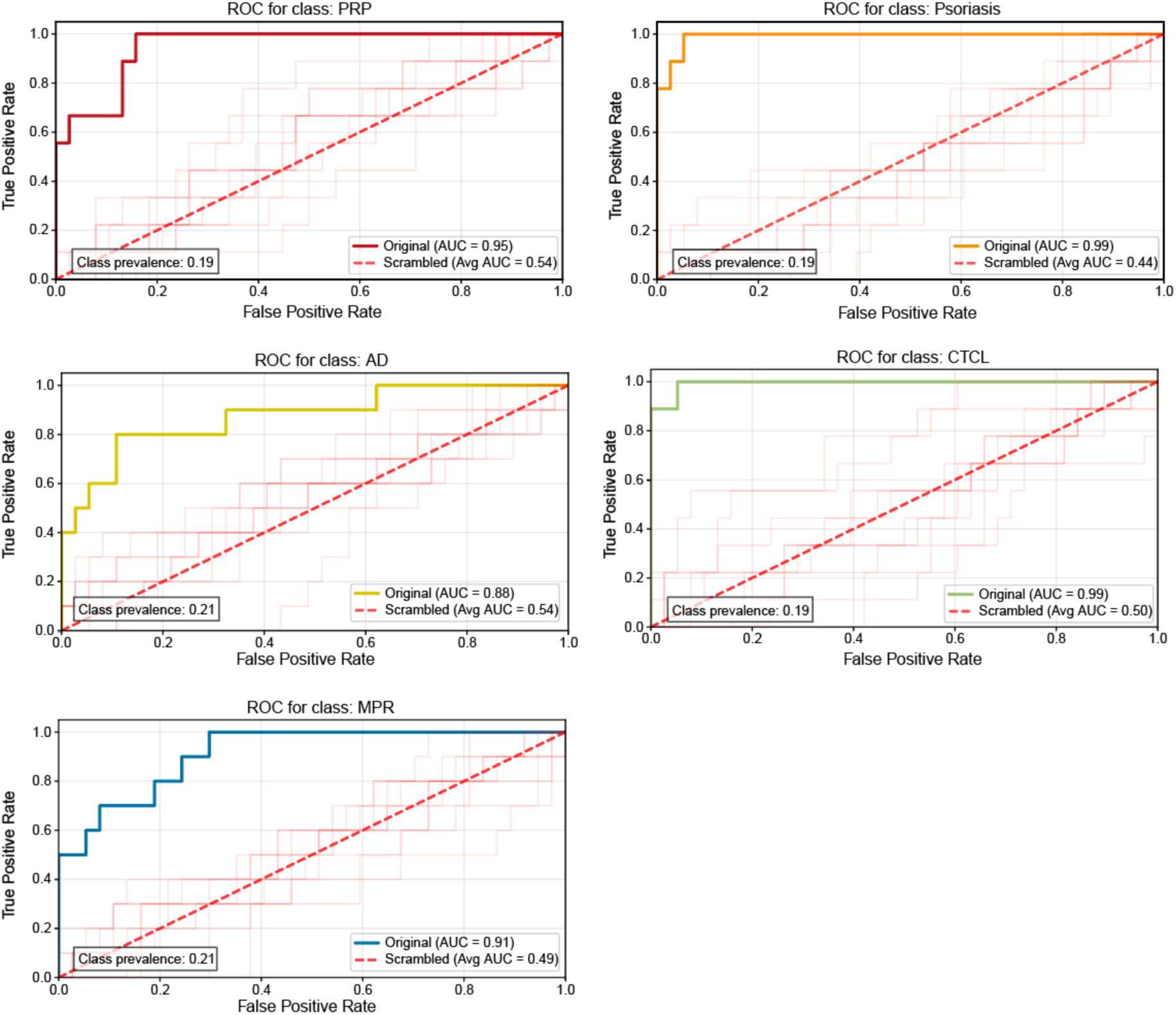
Scrambled ROC curves. ROC-AUC performance for the different erythrodermic diseases (pityriasis rubra pilaris, PRP; Psoriasis; atopic dermatitis, AD; cutaneous T-cell lymphoma, CTCL; maculopapular rash, MPR) by applying the multiclass classifier in comparison to scrambled average AUCs (n = 95 individuals).

